# Drug-induced decreased urine output predicts PDA closure in preterm neonates

**DOI:** 10.1101/2019.12.15.19014993

**Authors:** Malika Goel, Sourabh Dutta, Shiv Sajan Saini, Venkataseshan Sundaram

## Abstract

**Objective:** Prostaglandin inhibitors (PGI) are used to treat patent ductus arteriosus (PDA) of prematurity. PGIs often cause decrease in urine output (UO), the mechanism of which is like that of PDA closure. We hypothesized that PGI-induced decrease in UO predicts PDA closure.

**Design:** Prospective, cohort

**Setting:** Level III NICU

**Methods:** We prospectively enrolled 40 preterm neonates (≤34 weeks gestation) with clinical and/or echocardiographic hemodynamically significant PDA (hsPDA), being treated with Ibuprofen or Paracetamol. We measured UO, weight, total fluid intake (TFI) at baseline and daily until 72 h. We performed echocardiogram at baseline and daily until PDA closure or end of treatment. We compared “PDA-closed” and “PDA-open” groups for change in UO, weight and TFI from baseline.

**Results:** “PDA-closed” and “PDA-open” groups had 28 and 12 neonates respectively. Median (Q1, Q3) percent decrease in UO was greater in “PDA-closed” vs “PDA-open” group: from baseline to 0-24 h [-44.87% (−54.79%, +0.04%) vs −15.03% (−27.91%, +49.11%)]; to 24-48 hours [-40.85% (−52.81%, +14.45%) vs. −2.57% (−24.82%, +61.73%) and to 48-72 hours [-32.77% (−49.4%, +32.22%) vs. +20.74% (−6.88%, +98.39%). “PDA closed” group had significantly greater percent decrease in weight by 72-h. Mixed linear model showed that “group” and “time” were independently associated with UO; but “group*time” interaction and covariates (echocardiographic hsPDA, weight, gestation, postnatal age) were not. Decrease in UO of 27% and 17% by 24-48 h and 48-72 h respectively, best predicted PDA closure.

**Conclusions:** Transient decrease in UO after treating hsPDA with a PGI may predict successful closure of PDA.

## INTRODUCTION

Patent ductus arteriosus (PDA) is a common morbidity among premature neonates (1). The patency of the ductus is primarily maintained by hypoxemia and circulating prostaglandins (2). Increased sensitivity to prostaglandins in preterm infants causes persistence of PDA; hence, prostaglandin inhibition by drugs, such as indomethacin, ibuprofen and paracetamol, is a standard approach for the treatment of PDA (3).

Oliguria or deranged renal function is a frequent adverse effect of prostaglandin inhibitors (PGI) (4). Under conditions of decreased blood flow, prostaglandins cause afferent glomerular arteriolar dilatation. Cyclooxygenases I and II are required for prostaglandin synthesis. PGIs inhibit these enzymes, resulting in decreased renal blood flow and glomerular filtration.

Both indomethacin and ibuprofen increase the risk of oliguria compared to placebo (5, 6). A network meta-analysis on indomethacin, ibuprofen and paracetamol showed that all agents and modes of administration had a higher risk of oliguria compared to continuous intravenous infusion of Ibuprofen (3). In another systematic review, there was no difference in the incidence of oliguria between the paracetamol and ibuprofen(7). The above literature suggests that all the PGIs have the potential to affect urine output (UO)- to a greater or lesser extent. A far more common phenomenon is a transient decrease in the urine output (UO), not amounting to oliguria (8, 9).

The mechanism by which the PGIs effect PDA closure in preterm infants is the same mechanism by which they cause oliguria. We hypothesized that preterm infants whose ductal tissue shows greater sensitivity to PGIs are likely to show greater renal vascular sensitivity to the same. Thus, a transient decrease in UO immediately after starting treatment with PGIs may be associated with greater chances of PDA closure. Several predictors of PDA closure following treatment with PGIs have been described in literature (10-16). However, to the best of our knowledge, a transient decrease in UO from baseline as a predictor of PDA closure has never been described before. We were also cognizant that oliguria following medical treatment may increase intravascular volume, thus preventing the closure of a PDA.

We planned a prospective cohort study with the hypothesis that among preterm infants with a hemodynamically significant PDA (hsPDA) being treated with a PGI, a transient decline in UO, following drug administration, is a predictor of successful PDA closure.

## METHODS

We conducted this prospective cohort study in a level III neonatal intensive care unit in Northern India from January 2017 to May 2018. This is a university hospital that caters to a middle-class and lower middle-class population. There are approximately 6000 live births, 550 admissions to the NICU and 60 cases of hsPDA that are medically treated per annum. The unit has the facilities and expertise for bedside echocardiography. We use ibuprofen and paracetamol for treating PDA. The regimen for ibuprofen is 10 mg per kg followed by 2 doses of 5 mg per kg at 24-hour intervals; and for paracetamol 15 mg per kg per dose every 6 hours for 12 doses. The unit policy is to treat preterm PDA if it is clinically or echocardiographically hemodynamically significant. Diuretics are not used, unless there is pulmonary edema.

We screened all preterm newborn infants, admitted for the treatment of PDA. We included neonates delivered at ≤34 weeks of gestation who had an echocardiographically proven PDA with left-to-right shunt and who were being treated with a PGI (paracetamol or ibuprofen by any route). Our pre-enrolment exclusion criterion was administration of first dose beyond 14 days of life. Our post-enrolment exclusion criteria were subjects in whom the UO could not be reliably measured; who had an interrupted or incomplete course of the drug and in whom the status of the PDA could not be assessed echocardiographically by the end of the medication course (within 24 hours after the last dose). If the UO could not be measured for any reason or was mixed with stool in a diaper for more than 2 diapers in any 24-h period, we considered it to be an unreliable estimate. We obtained written informed parental consent before enrolling.

We recorded demographic variables, the time of closure of PDA, the medication used, details of fluid balance, evidence of sepsis prior to onset of PDA, and UO. We measured the catheter UO among those patients who were already catheterized prior to enrolment. We did not catheterize any patient solely for the study. We measured the UO of male neonates by the test tube method. Nurses aspirated the test tube at least 2-hourly and recorded the volume. We measured the UO of female neonates by 2-hourly diaper weighing. If the number of diapers where the urine was mixed with stool was ≤2 in a 24-h period, we imputed the urine volume in the soiled diaper by averaging the urine volume from the 3 previous wet diapers.

We measured blood urea and serum creatinine before starting the PGI. The total fluid intake (TFI) [in ml/kg/d] in a 24-hour period and the UO (in ml/kg/h) in a 12-hour period before the first dose of medication were taken as the baseline values. UO and TFI were measured for 24-hour periods starting from the first dose until 24 hours after the last dose. Neonates were weighed before starting treatment (baseline) and every 24 hours. Blood urea and serum creatinine were assayed at any time if clinically indicated, or once at the end of the medical treatment.

Investigators (V.S. and S.S.) with expertise in neonatal echocardiography, performed the baseline echocardiogram (MicroMaxx Portable Ultrasound Machine; SonoSite, Inc, Bothell, WA) and 24-hourly echocardiograms until PDA closure or 24 hours after the last dose of the medication, whichever was earlier. Echocardiographic closure of the PDA was the key outcome variable and percentage decrease in UO from baseline to 0-24 hours after the first dose of the medication was the key predictor variable.

We grouped patients as “PDA closed” or “PDA open”. We compared the groups for percentage change in UO from baseline to the 0-24-hour period, 24-48-h period and 48-72-h period after the first dose; percentage change in TFI from baseline to the above time periods; percentage change in weight from baseline to 24, 48 and 72 hours; proportion with oliguria (defined as <1 ml/kg/h) in the above time periods and maximum values of blood urea and creatinine. As neonates would have varying baseline UO’s, TFI’s and weights, we preferred to compare the percentage change from baseline rather than the actual values.

As there was no information available on our hypothesis in published literature, we were unable to perform a formal sample size calculation. The institute ethics committee approved the study protocol (INT/IEC/2017/547 dated 5-5-17).

### Statistical analysis

We tested numerical variables for normality of distribution by the Shapiro Wilk test and QQ plot and described them as mean (standard deviation) or median (1^st^, 3^rd^ quartile). We compared numerical variables between the groups by Student’s t-test or Mann-Whitney U test, depending upon distribution. We described categorical variables as percentages and compared them by Chi-square test or Fisher’s exact test, as appropriate.

Since the UO at sequential time periods was a repeated measurement, we also constructed a mixed linear model with “percentage change in urine output from baseline” as the outcome variable and Group (with “PDA open” as reference category), time, and an interaction term of Group*Time (with “PDA open*Time” as reference category) as the predictor variables. We adjusted for the covariates: birth weight, gestational age, age at medical treatment and presence of echocardiographic hsPDA.

We calculated the area under the receiver operator characteristics curves (AUC) with percentage change in UO from baseline to the 3 time periods as index tests and PDA closure as the reference standard. We determined cut-off values based upon the highest Youden’s index.

## RESULTS

We screened 55 preterm infants who met the inclusion criteria and were enrolled after informed parental consent. We excluded 9 infants as the urine output could not be reliably estimated and 6 infants who died before the course of medication could be completed. We analysed data of the remaining 40 eligible neonates. PDA closed in 28 of these infants and remained open in 12. There were 3 (10.7%) and 3 (16.7%) subjects who received ibuprofen in the “PDA closed” and “PDA open” groups respectively. The rest received paracetamol. In the above groups, 25 (89.3%) and 10 (83.3%) subjects respectively received the medication intravenously. In the “PDA closed” group, the median (1^st^, 3^rd^ quartile) duration until clinical closure of PDA was 2.5 days (2, 3) from the 1^st^ dose; and duration until echocardiographic closure was 3 days (2, 3) from the 1^st^ dose. No subject received diuretics.

Baseline characteristics were not significantly different between the 2 groups, except for PDA size which was larger in the “PDA open” group (p=0.01) [Table 1]. The decrease in urine output expressed as a percentage change from baseline was greater at all time periods in the “PDA closed” group compared to the “PDA open” group (Table 2). The difference fell short of being statistically significant in the 0-24-hour period (p=0.07) and was statistically significant in the 24-48 hour period [median decrease of 40.85% and 2.57% from baseline in two groups respectively, p=0.03] and in the 48-72 hour period [median decrease of 32.77% in the “PDA closed” group and increase of 20.74% in the “PDA open” group, p=0.02]. 2 subjects in each group developed oliguria and they recovered from oliguria beyond 72 hours. No patient had UO less than 0.5 ml/kg/h necessitating stoppage of the medication.

**Table 1.** Comparison of baseline parameters between “PDA closed” and “PDA open” groups.

| **Variable** | **PDA closed**<br>**(n= 28)** | **PDA open**<br>**(n= 12)** | **P**<br>**value** |
| --- | --- | --- | --- |
| Birth weight, grams, mean (SD) | 1116.67<br>(321.21) | 1116.67<br>(321.21) | 0.85 |
| Gestational age, weeks, mean (SD) | 29 (1.82) | 29 (2.61) | 0.91 |
| Males | 12 (42.9) | 6 (50%) | 0.68 |
| Apgar at 5 minutes, Median (Q1, Q3) | 8 (7,8) | 8 (7,8.75) | 0.81 |
| Age PDA detected, days, Median (Q1, Q3) | 4 (3, 4) | 4.5 (2.25, 6.75) | 0.21 |
| PDA size, mm, mean (SD) | 2.61 (0.69) | 3.22 (0.6) | 0.01 |
| PDA hemodynamically significant on echocardiography, n (%) | 27 (96.4) | 11 (91.7) | 0.52 |
| Age when 1<sup>st</sup> dose of medication was administered, days, median (Q1, Q3) | 4 (3, 4) | 4.5 (2.25, 7) | 0.33 |
| Blood urea in 24-h period prior to 1<sup>st</sup> dose, mg/dL, median (Q1, Q3) | 33 (25, 53) | 33 (23, 41) | 0.3 |
| Serum creatinine in 24-h period prior to 1<sup>st</sup> dose, mg/dL, median (Q1, Q3) | 0.7 (0.55, 0.9) | 0.4 (0.3, 0.7) | 0.06 |
| Baseline UO in 12-h period prior to 1<sup>st</sup> dose, ml/kg/h, median (Q1, Q3) | 3.50<br>(2.23, 4.65) | 3.11<br>(3.28, 1.25) | 0.21 |
| TFI in 24-h period prior to 1<sup>st</sup> dose, ml/kg/d, mean (SD) | 109.25<br>(21.28) | 120 (28.28) | 0.19 |
| Weight prior to 1<sup>st</sup> dose, g, mean (SD) | 1079.93<br>(357.05) | 1093.67<br>(318.572) | 0.69 |
| Culture +ve sepsis in 48-h prior to 1<sup>st</sup> dose, n (%) | 0 | 0 | NA |
| Clinical sepsis in 48-h period prior to 1<sup>st</sup> dose, n (%) | 21 (75) | 7 (58.3) | 0.54 |

PDA = Patent ductus arteriosus
SD= standard deviation
Q1, Q3= 1<sup>st</sup> quartile, 3<sup>rd</sup> quartile
UO= urine output
TFI= total fluid intake

**Table 2.**
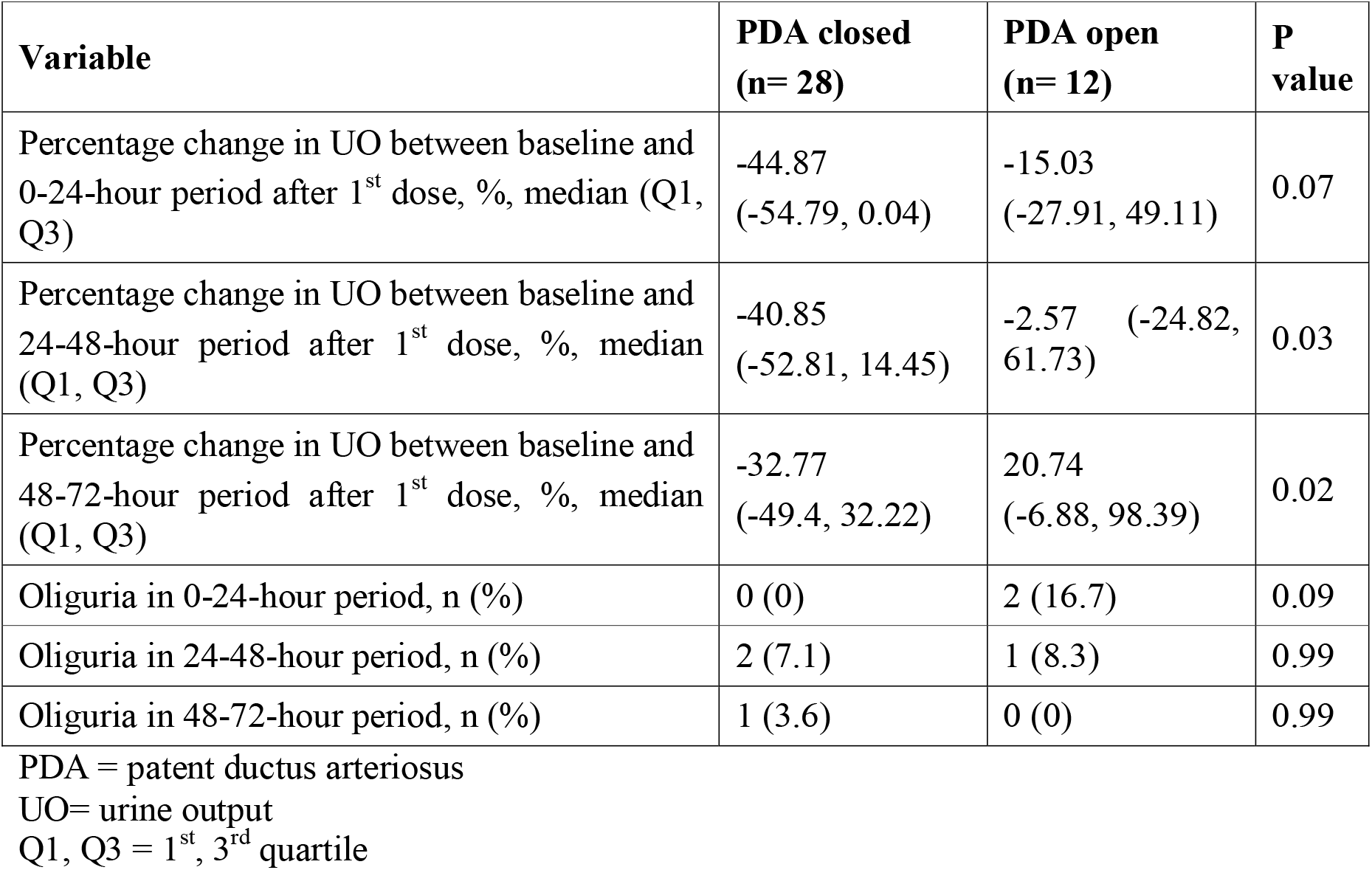
Comparison of urine output between “PDA closed” and “PDA open” groups.

| **Variable** | **PDA closed**<br>**(n= 28)** | **PDA open**<br>**(n= 12)** | **P**<br>**value** |
| --- | --- | --- | --- |
| Percentage change in UO between baseline and 0-24-hour period after 1<sup>st</sup> dose, %, median (Q1, Q3) | -44.87<br>(-54.79, 0.04) | -15.03<br>(-27.91, 49.11) | 0.07 |
| Percentage change in UO between baseline and 24-48-hour period after 1<sup>st</sup> dose, %, median (Q1, Q3) | -40.85<br>(-52.81, 14.45) | -2.57 (-24.82, 61.73) | 0.03 |
| Percentage change in UO between baseline and 48-72-hour period after 1<sup>st</sup> dose, %, median (Q1, Q3) | -32.77<br>(-49.4, 32.22) | 20.74<br>(-6.88, 98.39) | 0.02 |
| Oliguria in 0-24-hour period, n (%) | 0 (0) | 2 (16.7) | 0.09 |
| Oliguria in 24-48-hour period, n (%) | 2 (7.1) | 1 (8.3) | 0.99 |
| Oliguria in 48-72-hour period, n (%) | 1 (3.6) | 0 (0) | 0.99 |

PDA = patent ductus arteriosus
UO= urine output
Q1, Q3 = 1<sup>st</sup>, 3<sup>rd</sup> quartile

There were no statistically significant differences in the values of blood urea and serum creatinine (Table 3). There were no significant differences in the percentage change in weight from baseline up to 24 and 48 hours. However, by 72 hours there was a median ∼3% weight loss in the “PDA closed” group as opposed to 0.7% weight gain in the “PDA open” group (p=0.03). There were no significant differences between the groups with respect to the percentage change in TFI from baseline to any time point.

**Table 3.** Comparison of renal functions, weight and fluid intake between “PDA closed” and “PDA open” groups.

| **Variable** | **PDA closed**<br>**(n= 28)** | **PDA open**<br>**(n= 12)** | **P**<br>**value** |
| --- | --- | --- | --- |
| Maximum blood urea in observation period, mg/dL, median (Q1, Q3) | 31.5<br>(19.93, 52.5) | 31.5<br>(26.25, 42.75) | 0.86 |
| Maximum serum creatinine in observation period, mg/dL, median (Q1, Q3) | 0.5 (0.3, 0.65) | 0.4 (0.3, 0.48) | 0.16 |
| Percentage change in weight from baseline to 24 hours after 1<sup>st</sup> dose, %, median (Q1, Q3) | -0.59 (-1.95, 0) | 0 (-2.07, 2.5) | 0.22 |
| Percentage change in weight from baseline to 48 hours after 1<sup>st</sup> dose, %, median (Q1, Q3) | -0.91<br>(-2.52, 0.77) | 0 (-1.88, 6.18) | 0.08 |
| Percentage change in weight from baseline to 72 hours after 1<sup>st</sup> dose, %, median (Q1, Q3) | -3.05<br>(-5.57, -0.26) | 0.68<br>(-2.63, 6.29) | 0.03 |
| Percentage change in weight from baseline to 24 hours after last dose, %, median (Q1, Q3) | -2.93<br>(-5.93, 1.59) | 0.3<br>(-4.11, 6.1) | 0.26 |
| Percentage change in TFI from baseline to 0-24-hour period after 1<sup>st</sup> dose | 9.55 (0, 15.63) | 6.9 (0, 17.2) | 0.83 |
| Percentage change in TFI from baseline to 24-48-hour period after 1<sup>st</sup> dose | 17.42<br>(8.52, 37.5) | 15<br>(1.67, 34.77) | 0.36 |
| Percentage change in TFI from baseline to 48-72-hour period after 1<sup>st</sup> dose | 28.64<br>(15.71, 50) | 12.69<br>(0, 48.86) | 0.26 |
| Percentage change in TFI from baseline to 24-hour period after last dose | 43.14<br>(33.33, 69) | 25.36<br>(12.67, 69) | 0.35 |

PDA = patent ductus arteriosus
TFI= total fluid intake
Q1, Q3 = 1<sup>st</sup>, 3<sup>rd</sup> quartile

After adjustment for co-variates, at time zero, the intercept of the “PDA closed” group was 41.54 percentage points lower than that of “PDA open” [p=0.037] (Table 4). The slope of the “PDA open” regression line was positive (16.62), which meant that with passage of each 24-hour epoch, the UO increased by 16.62% in the “PDA open” group (p=0.015). The slope of the “PDA closed” regression line was negative compared to the reference category (−10.98%), which meant that with passage of each 24-hour epoch, there was a statistically nonsignificant decrease in UO by 10.98% compared to the “PDA open” line. None of the potential confounders that had been included as covariates in the mixed linear model had a statistically significant association with the outcome. A direct comparison of the estimated marginal mean values of the percentage change in UO from baseline between the “PDA closed” and “PDA open” groups showed a statistically significant difference (p=0.009). At the average values of all the covariates, the percentage decline in UO in the “PDA closed” group was 14.69%, whereas the UO increased in the “PDA open” group by 37.84%.

**Table 4.** Mixed linear model to predict “percentage change in urine output from baseline”, adjusted for baseline covariates.

| **Fixed Effects Estimates** |  |  |  |  |
| --- | --- | --- | --- | --- |
| *Parameter* | *Coefficient* | *Lower bound of 95% CI* | *Upper bound of 95% CI* | *P value* |
| Intercept | 40.47 | -251.37 | 332.31 | 0.78 |
| Group PDA closed<br>PDA open | -41.54 | -80.45 | -2.63 | 0.037 |
| Time | 16.62 | 3.37 | 29.87 | 0.015 |
| Interaction<br>(Group PDA closed) x<br>Time<br>(Group PDA open) x<br>Time | -10.98 | -26.83 | 4.85 | 0.17 |
| hsPDA on echo No<br>Yes | 14.94 | -62.11 | 91.99 | 0.7 |
| Birth weight in grams | 0.02 | -0.05 | 0.09 | 0.52 |
| Gestational age in weeks | -0.95 | -12.32 | 10.41 | 0.87 |
| Postnatal age at 1<sup>st</sup> dose | -5.49 | -13.05 | 2.07 | 0.15 |
| **Covariance parameters** |  |  |  |  |
| *Variance of* | *Estimate* | *Lower bound of 95% CI* | *Upper bound of 95% CI* | *P value* |
| Intercept | 4062.98 | 2568.18 | 6427.83 | <0.001 |
| Slope | 9899.41 | 6333.66 | 15,472.63 | <0.001 |
| Interaction of slope and intercept | 5521.95 | 2833.73 | 8210.17 | <0.001 |
| **Estimated marginal means of the groups and comparison between groups** |  |  |  |  |
| *Group* | *EMM\** | *Lower bound of 95% CI of EMM* | *Upper bound of 95% CI of EMM* | *P value of mean difference* |
| PDA closed | -14.69 | -55.69 | 26.31 | 0.009 |
| PDA open | 37.84 | -7.85 | 83.54 |  |

\*Covariates appearing in the model were evaluated at the following average values: Time =1 (ie 24-48-hours), birth weight= 1120.25 grams, gestational age= 29.03 weeks, age at the 1<sup>st</sup> dose of medication= 4.33 days
PDA = patent ductus arteriosus
CI = confidence interval
EMM = estimated marginal mean

When the percentage change in UO from baseline was used as a prognostic test to predict PDA closure, it had statistically significant but clinically modest AUC at 24-48 hours and 48-72 hours (Table 5). The AUC at 0-24 hours fell short of statistical significance (p=0.07). Based on these ROC curves, the best trade-off between sensitivity and specificity was achieved at a percentage decline in UO of approximately 34%, 27% and 17% in the 3 time periods respectively.

**Table 5.**
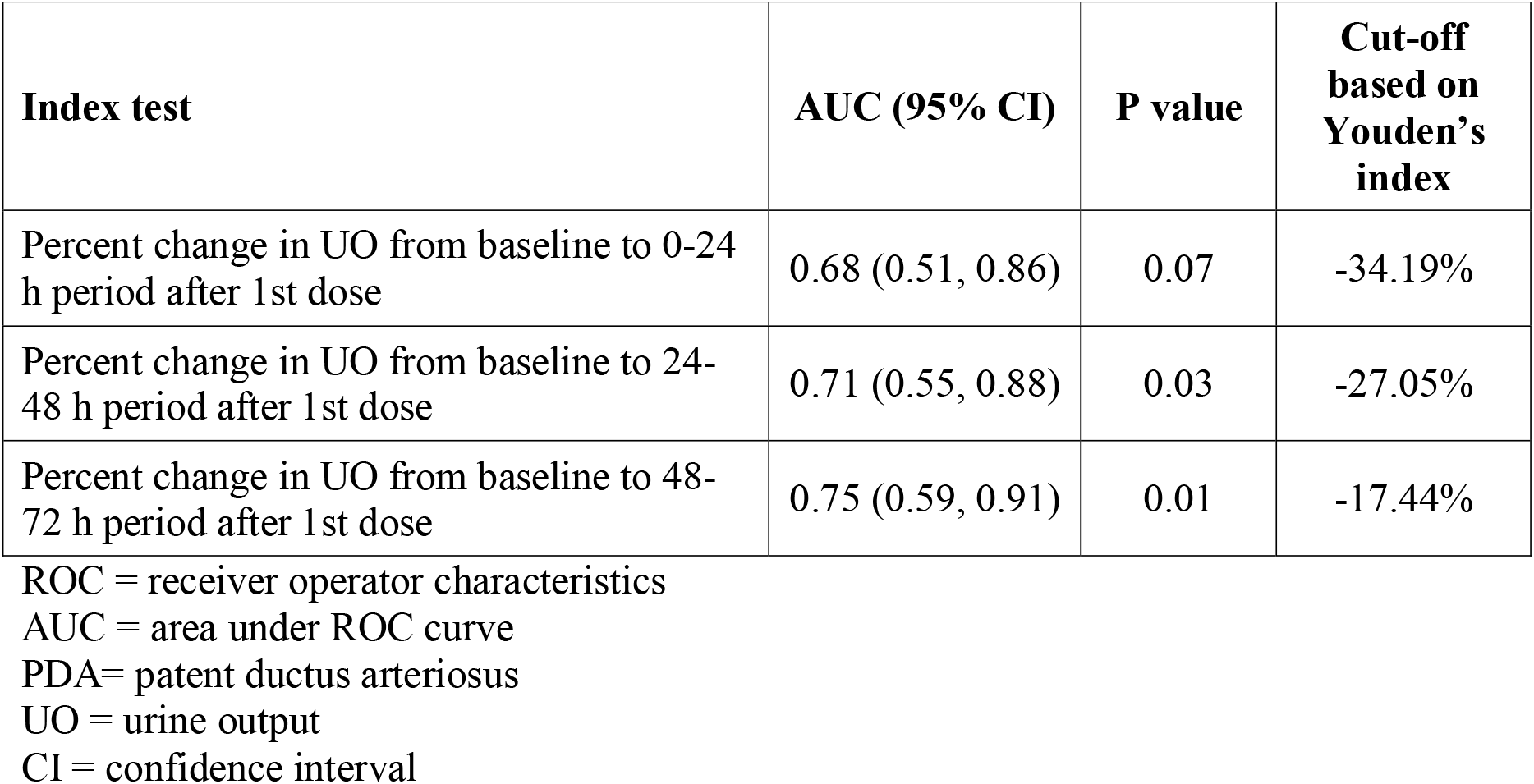
ROC curves and cut-offs based on Youden’s index for PDA closure as the outcome.

## DISCUSSION

The results of our study support our hypothesis that neonates with greater decrease in urine output from baseline following the administration of a PGI are more likely to have PDA closure. When observed over a 72-hour period following the start of medical treatment, there is a progressive gap between the change in UO from baseline in the “PDA closed” and “PDA open” groups. A 27% decline in UO by 48 hours after the first dose and a 17% decline by 72 hours after the first dose have the highest ability to predict PDA closure.

To the best of our knowledge, there are no other studies with which we can compare the results of our study. The literature is replete with studies regarding the incidence of decrease in urine output and renal dysfunction following the use of various PGIs for the treatment of PDA in preterm infants. However, none have evaluated a transient decrease in urine output as a clinical predictor for PDA closure.

The “PDA closed” and “PDA open” groups were comparable before the start of medication. The only difference was higher average PDA size among neonates whose PDA remained open, which is expected. Although a decrease in urine output was observed in most subjects, it was reassuring that oliguria (<1 ml/kg/h) was rarely observed, and when observed, it did not persist.

Before starting the study, we were also conscious of the possibility that decreased UO could result in volume overload and prevent PDA closure. However, our results do not support this possibility. The “PDA closed” group not only had significantly greater decrease in UO, but also significantly greater decrease in weight from baseline. This happened despite there being no significant differences in the TFI between the groups. This indicates that, while the decrease in UO is associated with closure of the PDA, there is no significant retention of fluid to cause weight gain. On the contrary, the closure of the PDA improves hemodynamics, resulting in less fluid retention and hence weight loss.

Since the UO at various time points are not independent data, we performed a mixed linear model. Our data did not meet the statistical assumptions required for performing a two-way repeated measures ANOVA. The mixed linear model reaffirmed that PDA closure had an independent relationship with percentage change in UO from baseline, after adjusting for co-variates.

Decrease in UO and closure of PDA appear to be epiphenomena, explained by a common underlying process of prostaglandin inhibition. The decrease in UO in the “PDA closed” group was apparent within the first 24 hours; whereas, the duration until echocardiographic closure had a median (1^st^, 3^rd^ quartile) of 3 days (2, 3). On average, the decrease in UO preceded the closure of the PDA and could serve as a bedside prognostic factor. Although hour-specific UO was recorded by the nurses, unfortunately, for the purpose of this research study we only recorded the UO in ml/kg/h for 24-hour periods in the case report forms. Hence, we are unable to comment whether the hour-wise decline in UO preceded ductal closure in individual patients.

The major limitation of the study was a small sample size. Hence, we were unable to include a larger number of covariates in the mixed linear model. We were also unable to split the sample randomly into a derivation cohort and validation cohort to test the validity of the ROC curves and the cut-offs based on the Youden index. However, despite the limitation of sample size, we were able to demonstrate findings that support our novel hypothesis. Larger studies are required to confirm our findings and to validate the cut-off values of percentage decrease in UO, so that the cut-off values could serve as a clinical tool during patient management.

The results of our study are generalisable to settings where paracetamol and ibuprofen are used for management of either clinical or echocardiographically significant PDA. Our study population had a mean birth weight of 29 weeks. The results may not necessarily be applicable to preterm infants more immature than this.

## Data Availability

Data can be made available on written request to the corresponding author, if the purpose is to use it for an individual patient data meta-analysis.

## ACKNOWLEDGEMENTS

None

## What is already known this topic

1. Prostaglandin inhibitors, such as paracetamol, ibuprofen and indomethacin, are used for the treatment of patent ductus arteriosus (PDA) of prematurity
2. These medications frequently cause decrease in urine output (UO), at times amounting to oliguria, mediated through prostaglandin inhibition in renal vasculature
3. Decrease in UO has never been evaluated as a predictor of PDA closure.

## What this study adds

1. Decrease in UO from baseline to 24 hours, 48 hours and 72 hours after start of medication predicts PDA closure.
2. Percent change in UO is independently associated with PDA closure after adjusting for important covariates
3. The decrease in UO is associated with weight loss rather than weight gain, with total fluid intake being similar

